# Factors influencing English general practitioners’ referrals to specialist sleep services: a qualitative study using the COM-B model

**DOI:** 10.1101/2025.10.26.25338808

**Authors:** Martina Sykorova, Frederick van Someren, Kristin Veighey, Ellen Nolte, Charlotte Warren-Gash, Michelle A. Miller, Sofia H. Eriksson, Ian E. Smith, Helen Strongman

## Abstract

**Title:** Factors influencing English general practitioners’ referrals to specialist sleep services: a qualitative study using the COM-B model

**Objectives:** This study explored the factors influencing access to sleep services for individuals with symptoms of OSA and narcolepsy, from the perspective of general practitioners (GP).

**Methods:** A qualitative interview study was conducted with GPs within three areas of England: South London, East Midlands or South West England to explore their views on factors influencing referrals to specialist sleep services. The semi-structured interviews were conducted between November 2024 and April 2025 using an interview guide informed by published research and the COM-B model of behaviour change; this model proposed that Capability (C), Opportunity (O), and Motivation (M) are needed for behaviour (B) change to occur. Data were analysed using exploratory thematic analysis informed by the COM-B model using an iterative approach.

**Results:** We conducted 31 interviews, mostly online, with one conducted face-to-face. Our data suggest that the most important factors shaping referral to sleep services are limited capacity of NHS sleep services, limited referral pathways for narcolepsy, inflexible referral pathways for OSA, and limited knowledge of narcolepsy.

**Conclusions:** This qualitative study with GPs in England highlights that, although sleep disorders are a common concern, the current healthcare system provides limited support for GPs in managing these conditions. Fundamental sleep medicine service reforms are needed to improve referral pathways. These reforms should be guided by data-driven research that assesses current services in relation to population health needs and evaluates the potential health and economic benefits of expanding service capacity.

## Introduction

Sleep is a fundamental determinant of human health, yet this has historically been under-recognised in medical practice. Sleep disorders are increasingly prevalent in the United Kingdom (UK) and worldwide, causing significant morbidity such as excessive daytime sleepiness (EDS) and substantially impaired quality of life. Sleepiness accounts for approximately 20% of all road collisions (British Lung Foundation, 2015), underscoring the importance of early diagnosis and treatment of EDS. Prior to their formal classification in 1990 (Thorpy, 1990), sleep disorders were largely regarded as consequences of other medical conditions and lifestyle factors. In contrast, this classification recognised primary sleep disorders of respiratory and neurological aetiology (e.g. Obstructive Sleep Apnoea (OSA) and narcolepsy, respectively). The standardized diagnostic prevalence in England is estimated at 1.4% for OSA (approximately 622,528 individuals) and 0.02% for narcolepsy (approximately 11,307 individuals) (Strongman et al., 2025). Despite increases in prevalence overtime, this is considerably less than symptomatic prevalence estimates of 0.047% for narcolepsy (Ohayon et al., 2002) and 4.8% for OSA (Benjafield et al., 2019). . Treating all currently untreated individuals with OSA could save the National Health Service (NHS) £28 million (British Lung Foundation, 2015). OSA and narcolepsy are typically diagnosed and treated by specialist sleep clinicians working individually or in umbrella services often centred in respiratory departments. In England, access to specialist services is coordinated by general practitioners (GP)who facilitate onward referrals to secondary care.

In a scoping review of qualitative evidence (Sýkorová et al., 2025), several factors influencing referrals to specialist sleep services internationally were identified. These included, a lack of role clarity, challenging referral processes, limited referral options, clinician knowledge of sleep disorders, funding arrangements, and attitudes towards treatment. To our knowledge there are no qualitative studies investigating factors influencing referrals to sleep centres in the UK or England. However, recently NHS England commissioned Getting it Right the First Time (GIRFT) respiratory report and British Sleep Society (BSS) Optimal Sleep Pathway report, both of which identified persistent barriers to delivering high-quality and equitable specialist sleep services and recommended optimal pathways for diagnosis and treatment (Allen, 2021; Read et al., 2024). These reports drew on expert clinical opinion, focus groups with healthcare professionals involved in the delivery of sleep medicine, questionnaires with NHS trusts, and limited routinely available hospital activity data. Sleep is not included in the GIRFT neurology report (Fuller, 2021).

As GPs were not consulted in the development of either the GIRFT respiratory or BSS Optimal Sleep pathway report, it is important to compare their perspectives with the recommendations in these reports. By capturing views from GPs working in areas with varying disease prevalence and differing access to specialist sleep services, this study aims to identify the factors that shape referral practices. These insights are essential for understanding contextual barriers and opportunities to support timely and appropriate referrals.

## Participants and methods

### Study design

This qualitative study used semi-structured interviews with GPs. The findings were analysed through the theoretical lens of the COM-B model of behaviour change (Michie et al., 2011), which suggests that behaviour change requires three elements: capability (C) to perform the behaviour, opportunity (O) to carry it out, and motivation (M) to enact it. This approach allowed us to systematically interpret GPs’ perspectives and identify potential influencing factors to behaviour change. We followed the consolidated criteria for reporting qualitative research (COREQ) guidelines to ensure rigour and transparency (Tong et al., 2007) (see online supplement).

### Ethics approval

The study was approved by the Health Research Authority (24/HRA/3320) and the London School of Hygiene and Tropical Medicine’s Ethics Committee (31009).

### Patient and Public Involvement statement

Our Patient and Public Involvement group included six members with diverse sleep disorders. They actively helped to design the study, refine study materials, and interpret findings, ensuring the study addressed relevant research questions.

### Dissemination to participants and related patient and public communities

Participants were offered the opportunity to receive the final study report upon completion. Findings will also be disseminated through patient and relevant organisations such as Narcolepsy UK, the Sleep Apnoea Trust and the BSS to ensure broad reach and accessibility.

### Participant selection

We aimed to recruit up to 40 GPs who were currently practising or recently retired (within 12 months), with recruitment continuing until data saturation was reached. Prior research suggests at least twelve interviews are needed to attain data saturation (Braun and Clarke, 2013; Guest et al., 2006). Our target sample size was informed by similar studies on barriers to sleep referrals, which conducted 20–48 interviews (Donovan et al., 2022; Germain et al., 2023; Graco et al., 2019; Grivell et al., 2021; Haycock et al., 2021; Hayes et al., 2012).

We used purposive sampling to recruit GPs in three Integrated Care Boards (ICBs), statutory NHS bodies in England that commission health services for local populations within the area, in the following regions: South London; East Midlands; and South West England. Instead of listing specific ICB areas, we report the broader regions where each ICB is located to maintain the anonymity of our participants. We have additionally replaced names and places with a pseudonym to protect participants’ identity.

The selected ICBs have low, average, and high predicted prevalence of OSA (Steier et al., 2014), respectively, and conversely high, average, and low prescribing of high-cost narcolepsy drugs (van Someren et al., 2024) and access to sleep centres.

The study was advertised through Regional Research Development Networks (RRDNs), organisations funded by the Department of Health and Social Care to support regional research delivery, via email or on the organisation’s research page. Interested GPs contacted the researcher (MS) via email or through the RRDN. One GP declined participation due to time constraints; no participants withdrew consent during or after interviews. Potential participants were sent an email containing a consent form and participant information sheet explaining the purpose of the study including information about data protection and confidentiality. Potential participants were asked whether they had any questions about the study and whether they would be willing to participate in the interview. Prior to the interview taking place, willing participants were asked to sign the consent form and email it to the researcher, and were given another opportunity to ask questions about the study.

### Data collection

We used an interview guide (see online supplement) developed based on findings from a scoping review (Sýkorová et al., 2025) and the COM-B model for Behaviour Change (Michie et al., 2011). The draft interview guide was revised based on feedback from a GP practising in the East Midlands. The interview guide and study objectives were also discussed with the project’s Patient and Public (PPI) Involvement and clinical/scientific advisory groups and amended based on their feedback. Further, a pilot interview was performed with a GP practicing outside the included ICBs – and finalised following the first four interviews. Each participant was only interviewed once. We adopted a researcher triangulation strategy by using two researchers (MS and HS) in the process of collecting interviews although most (n=29) interviews were conducted by MS. The interviews were limited to one hour and were offered to be conducted online (via Microsoft Teams) or in person.

### Data analysis

Data were analysed in two iterative stages using NVivo15 (Lumivero) and Excel. First, a six-stage inductive thematic analysis (Braun and Clarke, 2006) was conducted to allow themes and factors to emerge from data. Second, these factors and sub-factors were mapped onto the COM-B model. Initial coding was conducted by one coder (MS); further analysis was completed collaboratively by 3 coders (MS, HS, FvS) who collaboratively refined codes, developed overarching themes, and assigned COM-B domains. Patient and Public Involvement group members also reviewed sub-set of codes, leading to further refinement. Participants were not asked to evaluate the findings.

We report both barriers and facilitators influencing referrals to specialist sleep services to present a comprehensive overview. We also examined ‘negative cases’ i.e. instances that contradicted or deviated from dominant patterns to enhance the credibility of our findings and provide a more nuanced understanding of GPs’ perspectives on this complex issue.

### Reflexivity statement

Although this qualitative study reflects GPs’ perspectives, the researchers’ roles in its design, data collection, and analysis must be acknowledged, as they contribute to knowledge construction. The interviewers (MS and HS) both have sleep disorders, though this was not disclosed to participants prior to the interview to avoid influencing responses. Both researchers aimed to remain objective and set aside their presuppositions and personal experiences.

Participants were informed of the study’s purpose via the Participant Information Sheet. MS is a female qualitative researcher with training in qualitative methods; HS, also female, is an academic epidemiologist focusing on sleep research with training and experience in qualitative methods. A third coder, FVS - a male medical doctor with experience in clinical neurology and neurology research-also contributed to analysis. All three coders approached the data with openness and a commitment to being guided by the evidence.

## Results

We interviewed 31 GPs from England. Of the 30 interviewed GPs, eight were practising in South London, twelve within East Midlands, and ten within South West England. Most GPs practiced within urban (n=14), followed by rural (n=6), semi-urban (n=6) and mixed area (n=4). Participants practiced as GPs between 1 and 30 years. A half of GPs (14/30) reported involvement in the care for an individual with narcolepsy at some point in their career. All but one interview were conducted online. Interview length ranged from 24 to 65 minutes. Participants’ characteristics are reported in online supplement.

### Context

We conducted interviews during November 2024 – March 2025, a period when GPs in England were engaged in collective action protesting government proposals to change general practice funding. As part of this action, GPs prioritized core services and reduced non-essential tasks, such as completing hospital referral forms that were not part of their contractual obligations. Instead, they provided referral letters with the information they considered necessary, which specialist services were contractually obliged to accept (GP17, ID10, ID8). In some cases, however, specialists rejected these referrals, and GPs in turn challenged those rejections (GP17).

It is therefore important to note that our interviews captured GPs’ experiences of referring patients to specialist sleep services at a time when they were collectively resisting additional administrative burdens, such as lengthy referral forms. While our participants were all based in England, referral pathways, access to sleep centres or specialists, and eligibility criteria varied between individual Integrated Care Board (ICB) areas.

### Key themes

We identified three key themes: (1) Healthcare organizational system and arrangements, (2) Relationship between professionals and organisations, and (3) Individual perception of sleep disorders. Table 1 summarised the themes and key factors and sub-factors including the description of each sub-factors in more depth including corresponding quotes. The assigned COM-B domains are reported under each factor and sub-factor.

**Table 1:** Summary of overarching theme 1 and key factors mapped onto the COM-B model.

| Factors<br>(*assigned COM-B domains) | Subfactors<br>(*assigned COM-B domains) | Current status as reported by GPs | Example quotes |
| --- | --- | --- | --- |
| **Theme: Healthcare organizational systems and arrangements** |  |  |  |
| **Capacity of NHS sleep services**<br>*\*Physical opportunity* | **Insufficient provision of sleep specialists for narcolepsy**<br>*\*Physical opportunity* | While GPs knew of local sleep services for OSA within the ICB, access to narcolepsy specialists was limited, especially in one South West England ICB, reflecting the national neurologist shortage at the time of data collection. | "Um, so I think with regards to the narcolepsy side of things, having a local unit, so not having to refer to Primrose City. But I think because it's such a small – uh, it – it's such a specialist or rare condition, I think that's most probably why we have to refer out [of ICB3 area], if that makes sense."<br>(GP7, SWE) |
|  | **Long waiting times for consultations with sleep specialists**<br>*\*Physical opportunity* | Long waiting times to see a sleep specialist for OSA and narcolepsy represented a barrier in all three included areas. | "I think the problem with the waiting time in neurology is quite a significant one. So, I think quite often people would be waiting for about 18 months, so that's why I wouldn't, if, if, if hypothetically speaking, tomorrow I see a patient and I think it's a patient with narcolepsy, I think I'm going to really have a thought with myself. Is there any significant chances that it's not narcolepsy, but it's something that actually needs to be an ED, in A&E?"<br>(ID25, SWE) |
| **Referral pathways and guidance**<br>*\*Physical opportunity* | **Limited availability of referral pathways for narcolepsy**<br>*\*Physical opportunity*<br>*\*Reflective motivation* | Most GPs were not aware of, and could not find, a referral pathway for narcolepsy. The absence of a referral pathway for narcolepsy meant that GPs were unsure of where to refer patients with suspected narcolepsy which in some cases resulted in | "No, with – yeah, with narcolepsy, it's a bit more, um, of an uncommon, uh, diagnosis to make. It would be useful to have some clear guidance on-on when to refer patients with suspected narcolepsy, and what to include in-in referral letters, et cetera. That would be helpful, yeah." (GP30, SL)<br>"You know, I-I think the thing with sort of OSA is patients-patients would |
|  |  | GPs having no other choice than referring to the local respiratory sleep specialist. | *probably be slightly higher risk if they're overweight or these sorts of – you know, if their BMI and stuff, whereas it may not always be the case with somebody with narcolepsy. So I'd-I'd imagine it would probably be a similar referral pathway, Martina. We-we don't have a specific one for query narcolepsy. So I imagine I'd probably end up doing it on the same pathway as [laughs] I suppose sleep apnoea because it otherwise how would – how would I get them through and assessed?" (GP12, EM)* |
|  | **Strict and inflexible referral requirements and criteria for suspected OSA**<br>*\*Physical opportunity* | GPs found referral proformas helpful as they guided GPs on relevant information that sleep specialists would find helpful, however, strict and inflexible criteria were perceived as potentially excluding eligible individuals with OSA and limiting GPs' opportunity for clinical judgement. | *"Because we're getting, I mean we're finding, you know, huge amounts of our referrals are now getting rejected because of, you know. The thing is patients are people, they don't fit into neat categories. Um, and yes, if somebody is, is a very, very clear bog standard case of this [OSA], that or the other, then they fulfil the criteria easily. But not everybody fits into criteria easily. I mean I've had a few people who don't have daytime sleepiness but yet have been witnessed to stop breathing multiple times during their sleep or even had it recorded by a partner." (GP11, EM)* |
|  | **Patient self-referral to sleep services**<br>*\*Physical opportunity*<br>*\*Reflective motivation* | GPs suggested that an option to self-refer (given that individuals with OSA achieved a certain score on the Epworth Sleepiness Scale) to specialist sleep services would save GP time and decrease waiting times to see a specialist. | *"Um, [pause] well, I wonder even if a patient, you know, could patient self- seek, would there be a certain cohort who could almost do a self ... you know, if they presented with, you know, snoring or feeling tired, you know, could they just then do an Epworth, if it scored a certain amount and take me out then?" (GP24, SWE)* |

**Table 2:**
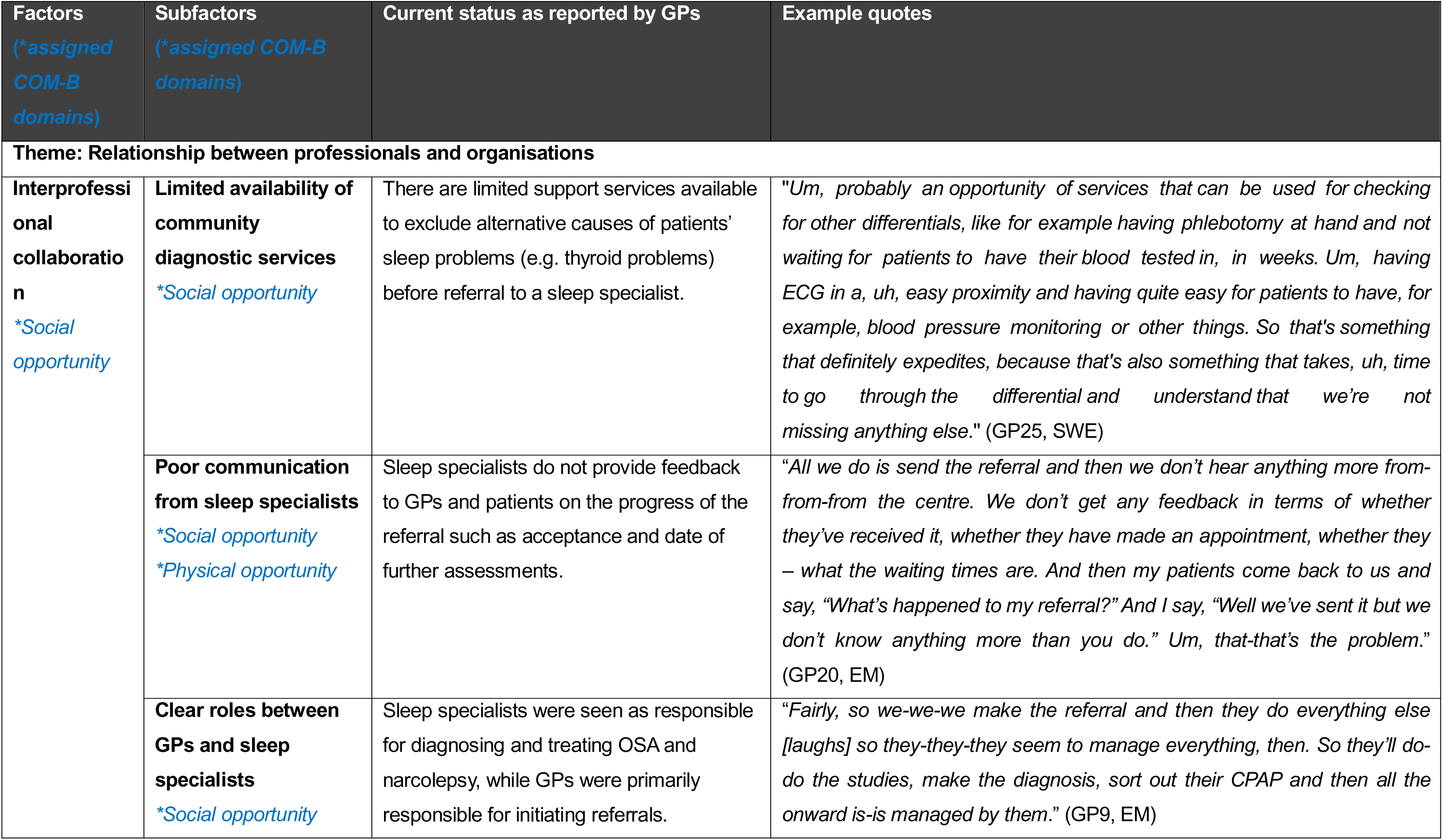
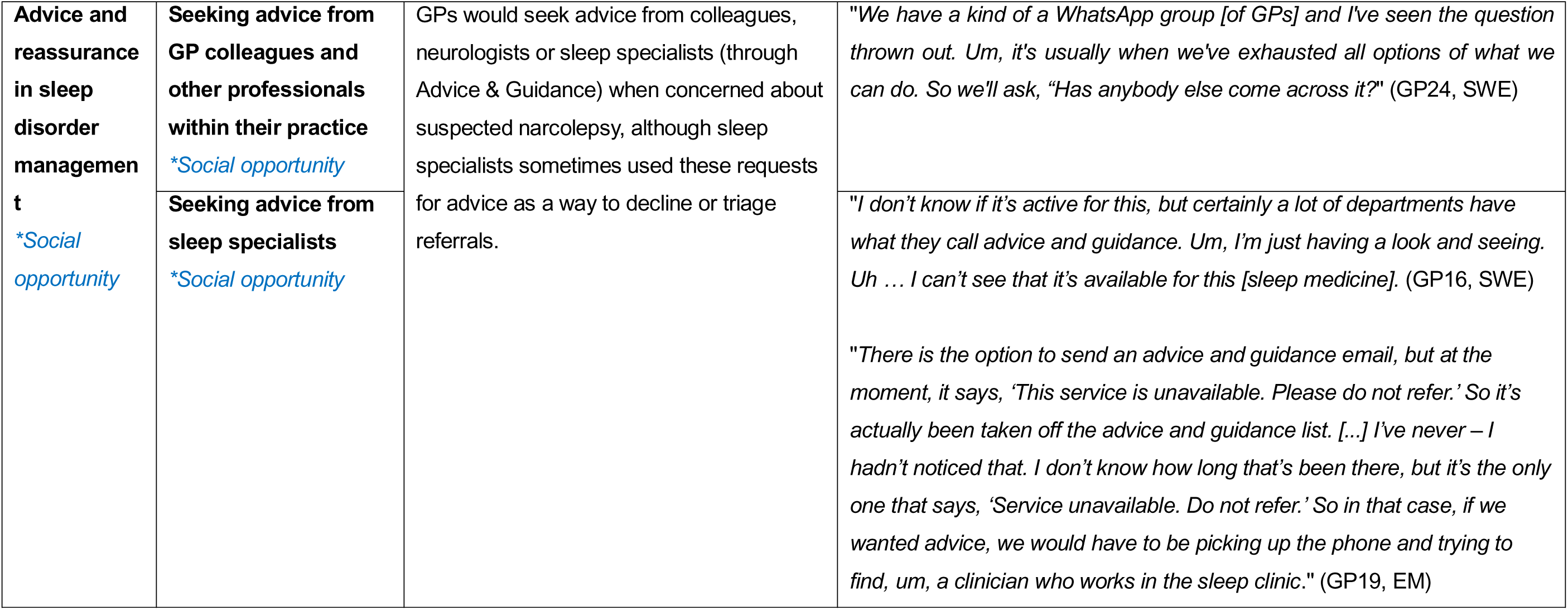
Summary of overarching theme 2 and key factors mapped onto the COM-B model.

**Table 3:**
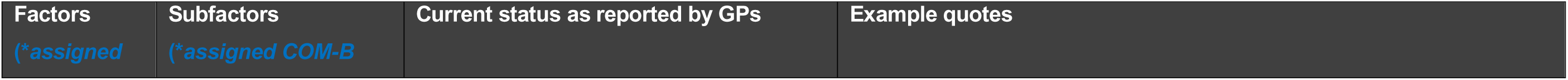
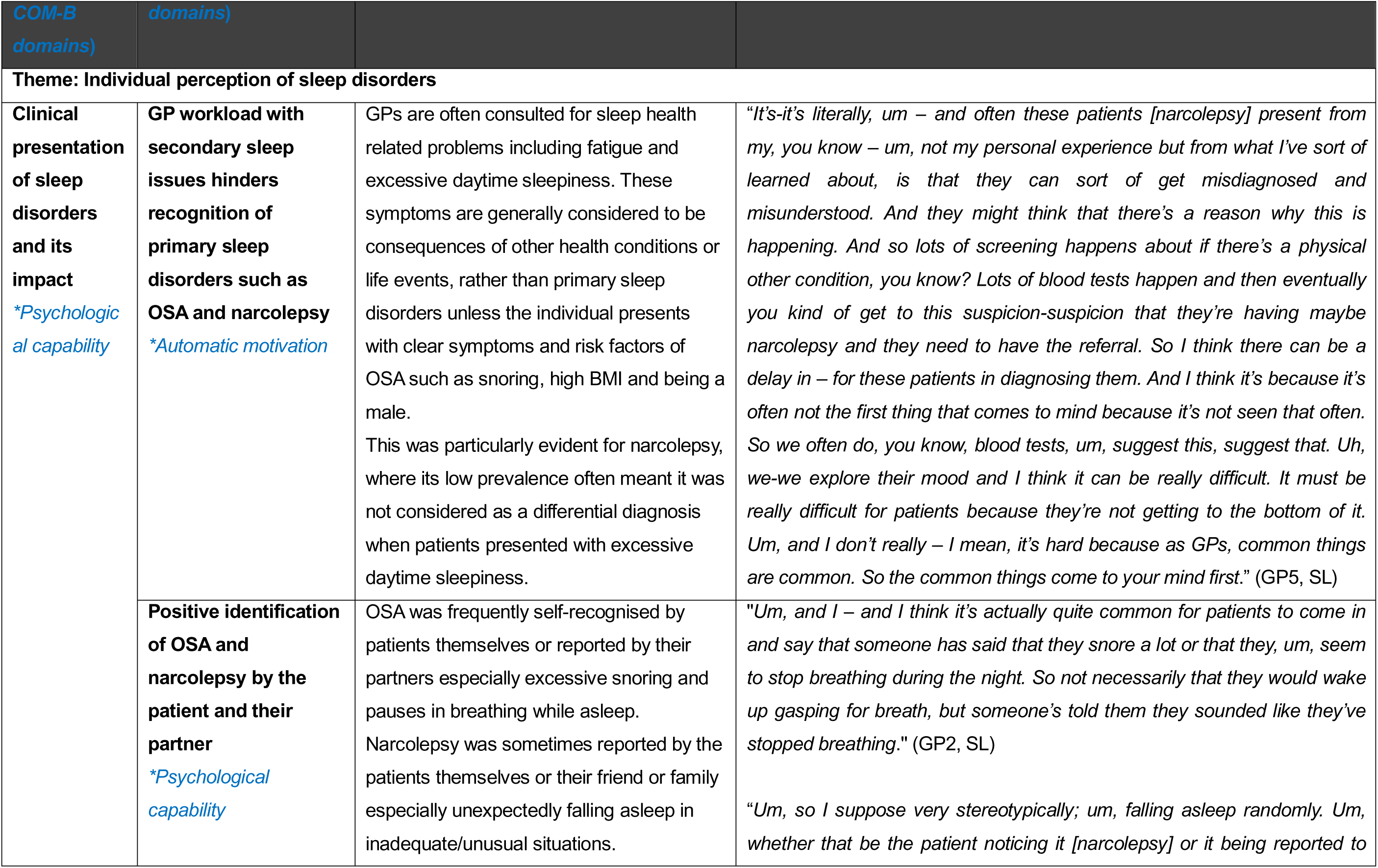
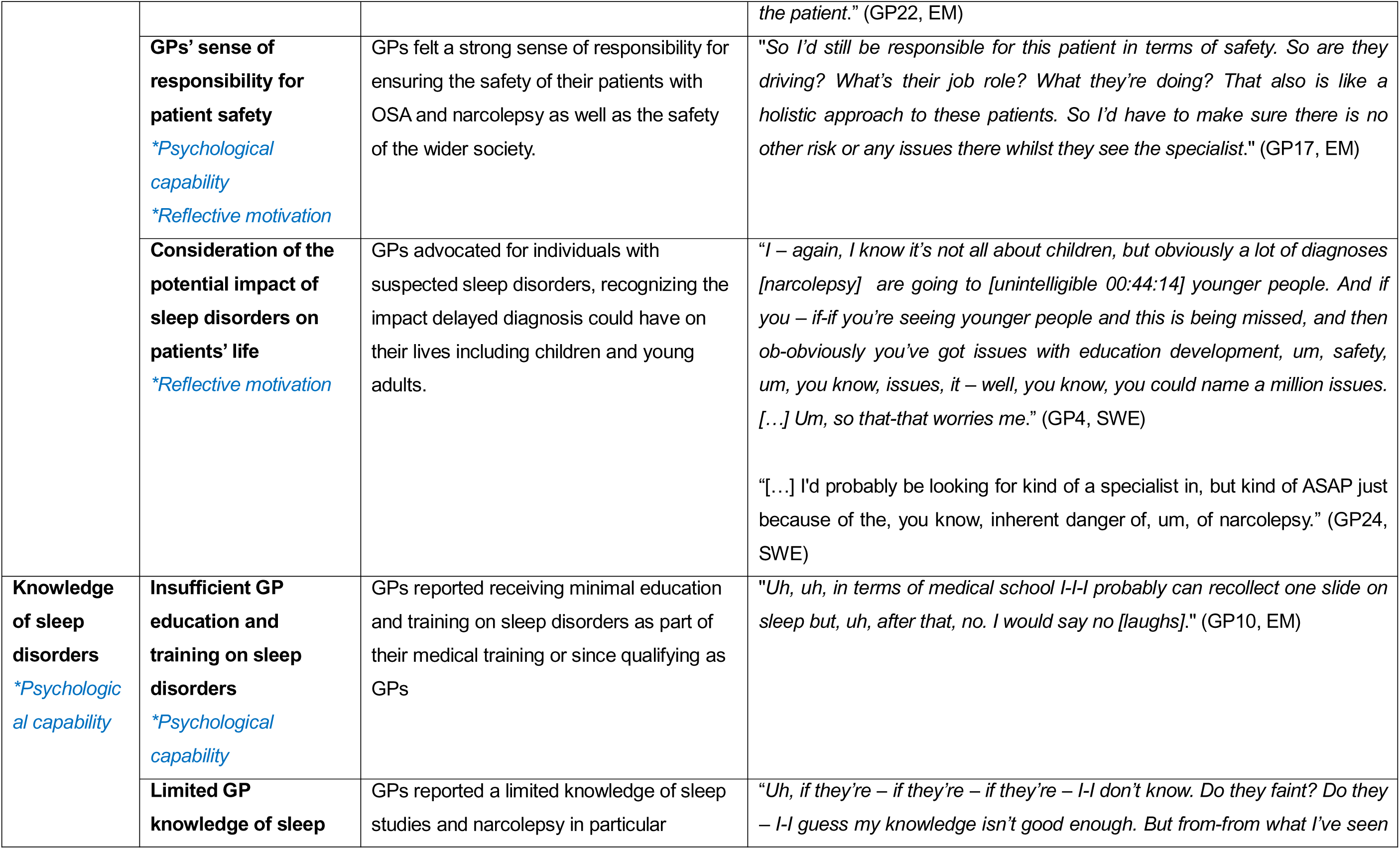
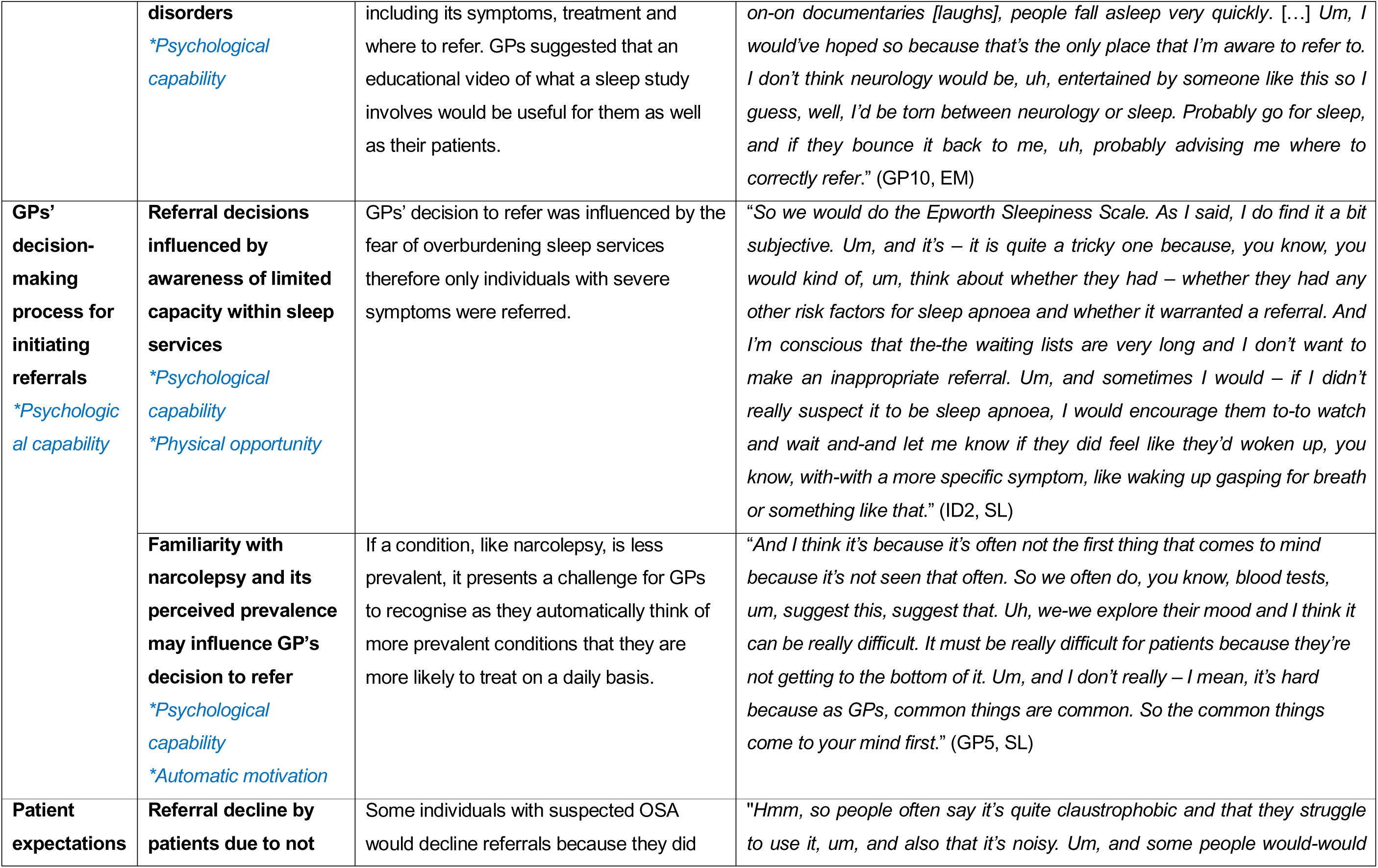
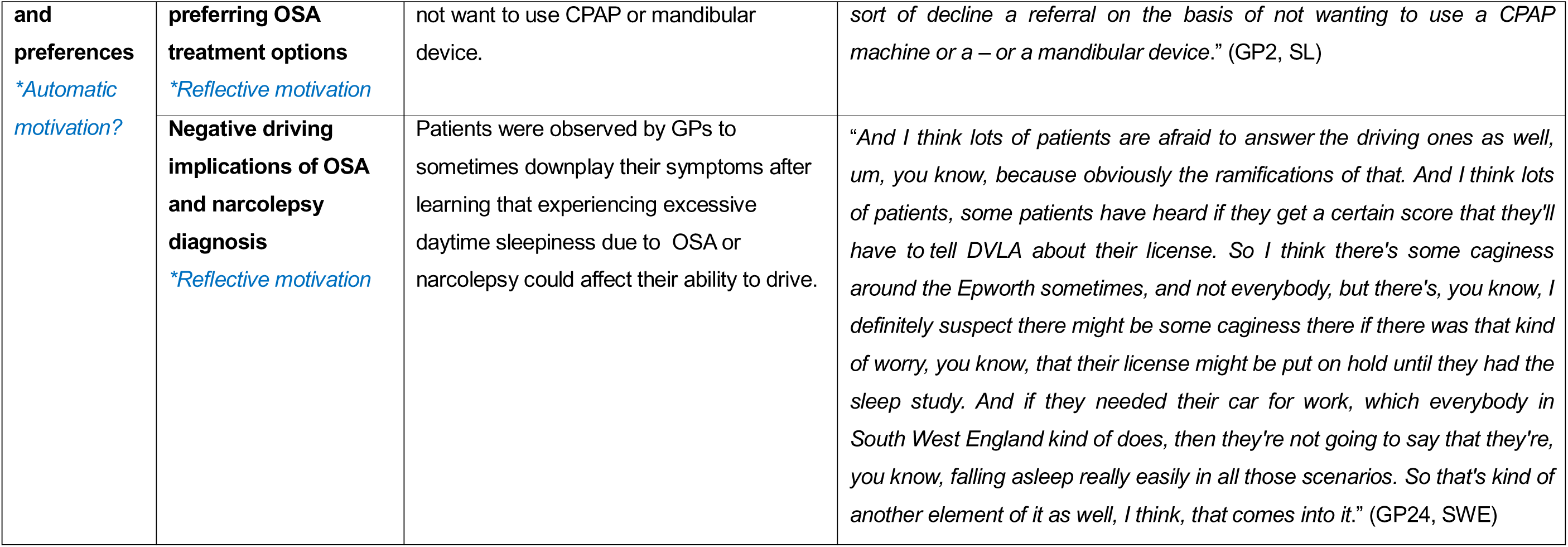
Summary of overarching theme 3 and key factors mapped onto the COM-B model.

| **Factors**<br>*(\*assigned* | **Subfactors**<br>*(\*assigned COM-B* | **Current status as reported by GPs** | **Example quotes** |
| --- | --- | --- | --- |

| COM-B domains) | domains) |  |  |
| --- | --- | --- | --- |
| **Theme: Individual perception of sleep disorders** |  |  |  |
| **Clinical presentation of sleep disorders and its impact**<br>*\*Psychological capability* | **GP workload with secondary sleep issues hinders recognition of primary sleep disorders such as OSA and narcolepsy**<br>*\*Automatic motivation* | GPs are often consulted for sleep health related problems including fatigue and excessive daytime sleepiness. These symptoms are generally considered to be consequences of other health conditions or life events, rather than primary sleep disorders unless the individual presents with clear symptoms and risk factors of OSA such as snoring, high BMI and being a male.<br>This was particularly evident for narcolepsy, where its low prevalence often meant it was not considered as a differential diagnosis when patients presented with excessive daytime sleepiness. | *"It's-it's literally, um – and often these patients [narcolepsy] present from my, you know – um, not my personal experience but from what I've sort of learned about, is that they can sort of get misdiagnosed and misunderstood. And they might think that there's a reason why this is happening. And so lots of screening happens about if there's a physical other condition, you know? Lots of blood tests happen and then eventually you kind of get to this suspicion-suspicion that they're having maybe narcolepsy and they need to have the referral. So I think there can be a delay in – for these patients in diagnosing them. And I think it's because it's often not the first thing that comes to mind because it's not seen that often. So we often do, you know, blood tests, um, suggest this, suggest that. Uh, we-we explore their mood and I think it can be really difficult. It must be really difficult for patients because they're not getting to the bottom of it. Um, and I don't really – I mean, it's hard because as GPs, common things are common. So the common things come to your mind first."* (GP5, SL) |
|  | **Positive identification of OSA and narcolepsy by the patient and their partner**<br>*\*Psychological capability* | OSA was frequently self-recognised by patients themselves or reported by their partners especially excessive snoring and pauses in breathing while asleep.<br>Narcolepsy was sometimes reported by the patients themselves or their friend or family especially unexpectedly falling asleep in inadequate/unusual situations. | *"Um, and I – and I think it's actually quite common for patients to come in and say that someone has said that they snore a lot or that they, um, seem to stop breathing during the night. So not necessarily that they would wake up gasping for breath, but someone's told them they sounded like they've stopped breathing."* (GP2, SL)<br>*"Um, so I suppose very stereotypically; um, falling asleep randomly. Um, whether that be the patient noticing it [narcolepsy] or it being reported to*<br>*the patient."* (GP22, EM) |
|  | **GPs' sense of responsibility for patient safety**<br>*\*Psychological capability*<br>*\*Reflective motivation* | GPs felt a strong sense of responsibility for ensuring the safety of their patients with OSA and narcolepsy as well as the safety of the wider society. | *"So I'd still be responsible for this patient in terms of safety. So are they driving? What's their job role? What they're doing? That also is like a holistic approach to these patients. So I'd have to make sure there is no other risk or any issues there whilst they see the specialist."* (GP17, EM) |
|  | **Consideration of the potential impact of sleep disorders on patients' life**<br>*\*Reflective motivation* | GPs advocated for individuals with suspected sleep disorders, recognizing the impact delayed diagnosis could have on their lives including children and young adults. | *"I – again, I know it's not all about children, but obviously a lot of diagnoses [narcolepsy] are going to [unintelligible 00:44:14] younger people. And if you – if-if you're seeing younger people and this is being missed, and then ob-obviously you've got issues with education development, um, safety, um, you know, issues, it – well, you know, you could name a million issues. [...] Um, so that-that worries me."* (GP4, SWE)<br>*"[...] I'd probably be looking for kind of a specialist in, but kind of ASAP just because of the, you know, inherent danger of, um, of narcolepsy."* (GP24, SWE) |
| **Knowledge of sleep disorders**<br>*\*Psychological capability* | **Insufficient GP education and training on sleep disorders**<br>*\*Psychological capability* | GPs reported receiving minimal education and training on sleep disorders as part of their medical training or since qualifying as GPs | *"Uh, uh, in terms of medical school I-I-I probably can recollect one slide on sleep but, uh, after that, no. I would say no [laughs]."* (GP10, EM) |
|  | **Limited GP knowledge of sleep** | GPs reported a limited knowledge of sleep studies and narcolepsy in particular | *"Uh, if they're – if they're – if they're – I-I don't know. Do they faint? Do they – I-I guess my knowledge isn't good enough. But from-from what I've seen* |
|  | **disorders**<br>*\*Psychological capability* | including its symptoms, treatment and where to refer. GPs suggested that an educational video of what a sleep study involves would be useful for them as well as their patients. | *on-on documentaries [laughs], people fall asleep very quickly. [...] Um, I would've hoped so because that's the only place that I'm aware to refer to. I don't think neurology would be, uh, entertained by someone like this so I guess, well, I'd be torn between neurology or sleep. Probably go for sleep, and if they bounce it back to me, uh, probably advising me where to correctly refer."* (GP10, EM) |
| **GPs' decision-making process for initiating referrals**<br>*\*Psychological capability* | **Referral decisions influenced by awareness of limited capacity within sleep services**<br>*\*Psychological capability*<br>*\*Physical opportunity* | GPs' decision to refer was influenced by the fear of overburdening sleep services therefore only individuals with severe symptoms were referred. | *"So we would do the Epworth Sleepiness Scale. As I said, I do find it a bit subjective. Um, and it's – it is quite a tricky one because, you know, you would kind of, um, think about whether they had – whether they had any other risk factors for sleep apnoea and whether it warranted a referral. And I'm conscious that the-the waiting lists are very long and I don't want to make an inappropriate referral. Um, and sometimes I would – if I didn't really suspect it to be sleep apnoea, I would encourage them to-to watch and wait and-and let me know if they did feel like they'd woken up, you know, with-with a more specific symptom, like waking up gasping for breath or something like that."* (ID2, SL) |
|  | **Familiarity with narcolepsy and its perceived prevalence may influence GP's decision to refer**<br>*\*Psychological capability*<br>*\*Automatic motivation* | If a condition, like narcolepsy, is less prevalent, it presents a challenge for GPs to recognise as they automatically think of more prevalent conditions that they are more likely to treat on a daily basis. | *"And I think it's because it's often not the first thing that comes to mind because it's not seen that often. So we often do, you know, blood tests, um, suggest this, suggest that. Uh, we-we explore their mood and I think it can be really difficult. It must be really difficult for patients because they're not getting to the bottom of it. Um, and I don't really – I mean, it's hard because as GPs, common things are common. So the common things come to your mind first."* (GP5, SL) |
| **Patient expectations** | **Referral decline by patients due to not** | Some individuals with suspected OSA would decline referrals because they did | *"Hmm, so people often say it's quite claustrophobic and that they struggle to use it, um, and also that it's noisy. Um, and some people would-would* |
| **and preferences**<br>*\*Automatic motivation?* | **preferring OSA treatment options**<br>*\*Reflective motivation* | not want to use CPAP or mandibular device. | *sort of decline a referral on the basis of not wanting to use a CPAP machine or a – or a mandibular device.” (GP2, SL)* |
|  | **Negative driving implications of OSA and narcolepsy diagnosis**<br>*\*Reflective motivation* | Patients were observed by GPs to sometimes downplay their symptoms after learning that experiencing excessive daytime sleepiness due to OSA or narcolepsy could affect their ability to drive. | *“And I think lots of patients are afraid to answer the driving ones as well, um, you know, because obviously the ramifications of that. And I think lots of patients, some patients have heard if they get a certain score that they'll have to tell DVLA about their license. So I think there's some caginess around the Epworth sometimes, and not everybody, but there's, you know, I definitely suspect there might be some caginess there if there was that kind of worry, you know, that their license might be put on hold until they had the sleep study. And if they needed their car for work, which everybody in South West England kind of does, then they're not going to say that they're, you know, falling asleep really easily in all those scenarios. So that's kind of another element of it as well, I think, that comes into it.” (GP24, SWE)* |

Additional barriers and facilitators were identified in the interviews that influence referrals to specialist sleep services, either for sleep disorders in general (not limited to OSA or narcolepsy) or for other conditions such as insomnia, can be found in the Online Supplement. We are reporting on factors that are specific to OSA and narcolepsy and relevant to the research question.

## Discussion

Our interview study of 30 GPs from three ICBs in England on factors affecting GPs’ referral practices to specialist sleep services identified three key themes: healthcare organisational system and arrangements, relationships between professionals and organisations and individual perceptions of sleep disorders. We found that while GPs are motivated to help people with suspected sleep disorders, they are overwhelmed by the volume of patients reporting sleep health problems, and work in a health system that lacks capacity for learning about, triaging, diagnosing and treating primary sleep disorders such as OSA and narcolepsy. Challenges vary substantially between ICBs and are greater for narcolepsy than OSA. We describe short-term opportunities to improve referral pathways and communication between sleep medicine specialists and GPs and consider the data-driven research that is needed to raise the profile of sleep medicine and motivate more fundamental reforms to NHS services.

One strength of the study lies in conducting an appropriate number of in-depth interviews with GPs from three English regions working in settings with varied access to sleep specialists. Data saturation was reached in each region when no new factors or insights emerged from subsequent interviews. Our study therefore incorporates a broad range of views reflecting the varied experiences of GPs across the NHS. To enhance the credibility of the analysis, multiple coders from different backgrounds reviewed the data, helping to mitigate potential biases.

However, only GPs practising in England were included; the devolved structure of healthcare governance in the UK and variation in healthcare provision internationally limits comparison across countries. Further, the absence of sleep specialist and patient perspectives constrained the study’s ability to fully elucidate the multifaceted nature of referral pathways. In this discussion, we use analysis and recommendations presented in clinician-led reports to provide the specialist perspective. Surveys of people with OSA and narcolepsy and their supporters provide the patient perspective.

A scoping review (Sýkorová et al., 2025) of international evidence showed that unclear roles and responsibilities, unclear referral pathways, referral to external networks, limited knowledge of sleep disorders, lack of funding and time constraints, sceptical views on treatment, and training and education on sleep disorders influence referrals to specialist sleep services. These factors largely align with our study. The more detailed evidence identified in the review was context specific often reflecting healthcare systems in Australia and the USA; no evidence was identified describing factors influencing referral for suspected narcolepsy.

The UK government and NHS England have prioritised the need to address waiting lists and regional variation in routine care (HM Government, 2024). To tackle this, NHS England commissioned GIRFT reports in multiple clinical areas including respiratory care (Allen, 2021) and neurology (Fuller, 2021). The neurology GIRFT report does not mention any neurological sleep disorders. The respiratory GIRFT report provides recommendations for the improvement of sleep medicine services following a clinically-led review encompassing questionnaire data, deep dive visits and routinely collected hospital activity data sources. Following this report, the BSS established a working group to review and provide recommendations on optimal sleep pathways. General practitioners were not consulted in these reviews and both reports predominantly focus on OSA.

Our findings on physical opportunity barriers to referral such as limited sleep service capacity and challenges with referral processes are consistent with both the GIRFT and BSS reports. We have described how these overlap with social opportunity barriers including poor communication between sleep specialists and GPs and the availability of community diagnostic and advice and guidance services. Both the GIRFT and BSS reports suggest that these physical and social opportunity barriers could be addressed with integrated services delivering the right tests and treatment at the right time and in the right place. This would involve more localised sleep apnoea diagnosis pathways and allow larger services to deliver more complex sleep services (e.g. narcolepsy diagnosis and treatment). To influence behaviour, current and future pathways would need to be clearly communicated to GPs as those participating in our study currently look to general neurologists or local respiratory sleep specialists when considering referrals for narcolepsy in areas without an established referral pathway. The GIRFT report calls for local sleep services in local hospitals that concentrate on confirming OSA diagnosis, initiating CPAP, and monitoring through technology-enabled CPAP. The BSS’s optimal sleep pathway would replace these local hospital services with community diagnostic hubs/centres or one stop clinics that co-ordinate home-based sleep studies supported by NICE clinical knowledge summaries, Specialist Advice and Guidance, a digital minimum dataset for sleep centre referrals, and potentially novel GP with Special Interest in Sleep roles. In common with GPs participating in our study, both reports recommend pre-referral testing and management as recommended by NHS England for patients with suspected chronic obstructive pulmonary disease, asthma, or heart failure (NHS England, 2023).

Specialists and GPs have a common psychological capability and reflective motivation to ensure rapid diagnosis especially in people with vigilance critical occupations and children and young adults (GPs only). This is in line with NICE guidelines for the diagnosis and management of sleep apnoea (NICE, 2021). However, GPs indicate that current inflexible digital referral forms may prevent referral of people with non-standard OSA, particularly those scoring below 12 on the Epworth Sleepiness Scale, despite NICE guidance acknowledging that not all individuals with OSA exhibit excessive daytime sleepiness.

In addition, reflective motivation is decreased when processes require GPs to convince patients of the benefits of CPAP prior to referral or patients downplay their symptoms due to fear of losing their driving license. Furthermore, there is not always an option to indicate urgency on referral forms and capacity of, and trust in, Specialist Advice and Guidance services needs to be increased as GPs believe that they are sometimes used to decline referrals (social opportunity). This, together with a fear of overburdening specialist services and feeling overwhelmed with consultations for sleep related problems including fatigue and excessive daytime sleepiness hinders the psychological capability and automatic motivation to refer. These psychological capability barriers are compounded by a lack of familiarity with narcolepsy and the perception that it is as an extremely rare condition. Some GPs noted self-identification of OSA and narcolepsy as a psychological capability facilitator to diagnosis and suggested that self-referral for OSA-diagnosis could reduce delays. This would overcome capability, opportunity, and motivation barriers notably in relation to a lack of training in sleep medicine from medical school onwards. This recommendation may underscore the impact of GP workload with secondary sleep issues on their automatic motivation to refer patients with symptoms suggestive of sleep disorders or to actively seek out opportunities for sleep medicine training.

THE GIRFT report suggests that in order to enact behaviour change, the motivation of commissioners to develop sleep services needs to be increased. To achieve this, data capture needs to be improved to allow trusts to model demand, measure the costs and benefits of delivering services, and ultimately negotiate with commissioners. Similarly, the BSS report discusses the need for adequate funding and analysis of Specialist Advice and Guidance activities; they suggest that local commissioners would be responsible for this. Service redevelopment will therefore require the profile of sleep medicine as a specialism to be increased – as the GPs in our study and international research (Sýkorová et al., 2025) suggest, this could be partly achieved with sleep medicine training from the undergraduate medical stage onwards (Romiszewski et al., 2020). As with Extended Role frameworks in areas such as dermatology and palliative care (RCGP, 2025), there is scope to develop an extended role for GPs with a special interest in sleep. Additionally, improved training and education on sleep disorders should also be available to other healthcare professionals as recommended by the Optimal Sleep Pathway (Read et al., 2024).

In the short term, commissioners and sleep specialists should ensure that there are documented referral and Advice and Guidance services for narcolepsy in all areas and that referral pathways for OSA allow urgency to be indicated and are in line with NICE guidance to avoid preventing referral of people whose quality of life and health is severely affected by OSA. Furthermore, people with symptoms suggestive of OSA and narcolepsy could be supported by patient organisations when presenting to GPs. A patient organisation survey of individuals with narcolepsy and their supporters suggested that less than a third were taken seriously when presenting to GPs (Narcolepsy UK, 2023). This survey identified similar social opportunity and psychological capability barriers to referral to our study including initial misdiagnosis, inappropriate referral to multiple specialists including general neurologists, and a reluctance to consider a narcolepsy diagnosis as it is rare. A survey of individuals with OSA indicated that 58% of individuals with OSA were referred to a sleep clinic after one visit (British Lung Foundation, 2014); this supports our finding that while GPs are more confident in recognising OSA than narcolepsy referral pathways could be improved. As GPs make the majority of referrals to specialist sleep services, GP representatives should be invited to contribute to sleep societies that develop care pathways.

Future research should also incorporate the perspectives of patients and sleep specialists (including paediatric specialists) and evaluate theory-informed interventions aimed at promoting equitable and efficient referral pathways for sleep disorders in England and the rest of the UK. Given that the areas with the highest predicted prevalence of OSA have the lowest density of sleep services (Steier et al., 2014), there is an urgent need to evaluate the service and pathways provision across the UK to reform sleep services. Moreover, as specialists in sleep centres manage a broad spectrum of conditions, future work should capture the perspectives of clinicians involved in treating sleep disorders beyond OSA and narcolepsy.

## Conclusions

This qualitative study with GPs in England highlights that, although sleep disorders are a common concern, the current healthcare system provides limited support for GPs in managing these conditions. Fundamental sleep medicine service reforms are needed to improve referral pathways. These reforms should be guided by data-driven research that assesses current services in relation to population health needs and evaluates the potential health and economic benefits of expanding service capacity.

## Data Availability

All data produced in the present study are available upon reasonable request to the authors

## Declaration of Generative AI and AI-assisted technologies in the writing process

During the preparation of this work the author used ChatGPT in order to improve clarity of the writing by checking grammar. After using this tool, the author reviewed and edited the content as needed and take full responsibility for the content of the publication.

## Acknowledgements

The authors would like to thank our Patient and Public Involvement group for their continuous support, guidance, and valuable contributions that helped shape this project.

## CRediT authorship contribution statement

**Martina Sykorova**: conceptualization, data curation, formal analysis, investigation, methodology, project administration, visualization, writing – original draft.

**Frederick van Someren**: formal analysis, methodology, writing – review and editing.

**Kristin Veighey:** supervision, writing – review and editing.

**Ellen Nolte**: supervision, writing – review and editing.

**Charlotte Warren-Gash:** supervision, writing – review and editing.

**Michelle A. Miller**: supervision, writing – review and editing.

**Sofia H. Eriksson**: supervision, writing – review and editing.

**Ian E. Smith**: supervision, writing – review and editing.

**Helen Strongman**: conceptualization, data curation, formal analysis, funding acquisition, investigation, methodology, supervision, writing – review and editing.

