## Supplementary material for "Factors influencing English general practitioners’ referrals to specialist sleep services: a qualitative study using the COM-B model": Online supplement

**Participants’ characteristics**

| **ID** | **Integrated Care Board area** | **Area** | **Years as a GP** | **Online/ face-to-face interview** | **Interview length** | **Encountered narcolepsy patient/s** |
| --- | --- | --- | --- | --- | --- | --- |
| GP1* | South London | Urban | 16-20 | Online | 00:31:51 | No |
| GP2 | South London | Urban | 6-10 | Online | 00:30:26 | No |
| GP3 | South London | Urban | 20+ | Face-to-face | 00:54:00 | Yes |
| GP4 | South West England | Rural | 0-5 | Online | 01:05:44 | Yes |
| GP5 | South London | Urban | 0-5 | Online | 00:28:41 | No |
| GP6 | South West England | Mixed | 16-20 | Online | 00:33:50 | Yes |
| GP7 | South West England | Urban | 0-5 | Online | 00:38:08 | Yes |
| GP8 | South West England | Urban | 0-5 | Online | 00:35:05 | Yes |
| GP9 | East Midlands | Urban | 20+ | Online | 00:32:17 | Yes |
| GP10 | East Midlands | Urban | 0-5 | Online | 00:38:53 | No |
| GP11 | East Midlands | Semi-urban | 16-20 | Online | 00:52:48 | Yes |
| GP12 | East Midlands | Semi-urban | 0-5 | Online | 00:41:38 | Yes |
| GP13 | South West England | Rural | 0-5 | Online | 00:37:34 | Yes |
| GP14 | East Midlands | Rural | 11-15 | Online | 00:45:31 | No |
| GP15 | South West England | Mixed | 11-15 | Online | 00:28:13 | No |
| GP16 | South West England | Rural | 16-20 | Online | 00:43:41 | No |
| GP17 | East Midlands | Mixed | 0-5 | Online | 00:43:00 | No |
| GP18 | East Midlands | Urban | 6-10 | Online | 00:39:53 | Yes |
| GP19 | East Midlands | Semi-urban | 11-15 | Online | 00:39:10 | No |
| GP20 | East Midlands | Semi-urban | 11-15 | Online | 00:47:49 | No |
| GP21 | South West England | Urban | 6-10 | Online | 00:40:02 | No |
| GP22 | East Midlands | Semi-urban | 11-15 | Online | 00:39:52 | No |
| GP23 | East Midlands | Rural | 6-10 | Online | 00:44:20 | No |
| GP24 | South West England | Urban | 0-5 | Online | 00:49:52 | No |
| GP25 | South West England | Mixed | 0-5 | Online | 00:50:34 | Yes |
| GP26 | South London | Urban | 6-10 | Online | 00:56:42 | Yes |
| GP27 | South London | Semi-urban | 0-5 | Online | 00:46:18 | No |
| GP28 | East Midlands | Rural | 20+ | Online | 00:44:53 | Yes |
| GP29 | South London | Urban | 0-5 | Online | 00:52:55 | No |
| GP30 | South London | Urban | 0-5 | Online | 00:46:11 | Yes |
| GP31 | South London | Urban | 16-20 | Online | 00:23:57 | No |

**Additional factors**

| **Other significant factors affecting referrals to sleep services that were reported by GPs but not relevant to our research question** | |
| --- | --- |
| **Factors** | **Corresponding quote** |
| Limited or no access to CBTi providers for insomnia | “*The only thing they [sleep centre] won’t see is insomnia. […] Not commissioned*.” (GP3, SL) |
| Private sleep providers issue especially in relation to shared care agreement and no follow up | “*But the problem is, as well, with things – like you talked about shared care agreements. The problem is, if a patient pays as a one-off to see a consultant and pays out of their own pocket, they’re quite likely not to go to the follow-ups because they have to pay for the follow-ups, and then that’s a problem with a shared care agreement or with equipment that needs maintaining. We see this a lot with conditions like ADHD, where it’s a very long waiting list to go and be seen and to get put on a drug that should be shared care, but then they’ll drop out of seeing the specialist because it’s costing them. But then we can’t keep prescribing. And they don’t understand that conflict, that we can only sign a prescription if you’re still seeing the consultant*.” (GP16, xx) |
| Untimely discharge from sleep services (OSA) which means that sometimes GPs need re-ref patients again (who then wait again for months) | “*Then they’re referred across to the, um, sleep nurses, who will – or the sleep physiologists, rather – who will then fit them with a machine and, uh, arrange follow-up. And then you will get to a point where they are, for example, discharged from the hospital, but yet they have a machine. And then sometimes you’ll see patients who come in because their machine is broken, for example, or they can’t fix it. And then you’re in a bit of limbo, because then you have to refer them back to – say they already have a diagnosis; they just need replacement materials. And they’ll be waiting 18 months for that, for example. Yes. It’s-it’s a very bizarre system. They don’t have any point of contact where patients can just dial a number and call for-for fixing of their equipment, which you would think would be common sense. Um, but they don’t*.” (GP18, xx) |
| GPs working in areas with a proportion of non-English speakers or speakers of limited English often require an interpreter which makes consultation appointments longer | “*We do, yes. So, we do quite a lot of interpreter appointments. I would say probably about 20% of my appointments are with an interpreter, so it’s quite high. […]. So I would say that consultation probably does take a bit longer, particularly if you're doing it via an interpreter as well*.” (GP29, SL) |
| Some sleep specialists lack knowledge of primary care since they spent all their working lives in specialist services which impacts sleep specialists’ understanding of what can be accomplished in primary care – this could be addressed by a placement in primary care | “*I think they’re probably, um … there is always a bit of a feeling that some specialists don’t understand primary care. They’ve never worked in primary care and they don’t understand where the – the limit – where the cut-off is between what’s specialist and what’s primary care. Because they work in a very narrow field. Um, and I think the impression is that neurology is probably one of the most distant from primary care; that they have the least understanding of what can and can’t be done in primary care*.” (GP16, xx) |
| **Barriers to referrals in general in general practice** | |
| Limited time and resources in general practice – GPs feel that appointments are too short | “*I-I-I – in-in-in a nice way, Martina, is that a serious question? [Laughs] Uh, no, absolutely not. I mean, it’s disgusting really, to be honest with you, the-the-the lack of time that we have in general practice. It’s-it’s disgraceful, uh [laughs], and I don’t – I don’t use those words lightly, and it adds-adds stress. It’s-it’s a very, very difficult place in which to work and, yeah, uh, patients sometimes may – I dunno, not that they say this to me – but I know some patients feel rushed and feel, you know? It’s just it’s horrible. It really is, so no, absolutely not. Do we have more time – would-would I like, um, the Health Secretary, um, to watch this video and-and-and-and-and [laughs] watch other people, sit and listen to what they say? Absolutely. Do we have enough time? No, no*.” (GP12, EM) |
| For certain conditions, GPs need to see patients several times to decide whether and where to refer | “*And for me, say, I know we’re not talking about urinary symptoms here. But there’s one for lower-lower urinary tract symptoms where I’ll get the patient to go away and complete the questionnaire because they will – they will need a second appointment anyway. That’s for urinary symptoms*.” (GP12, EM) |

**Interview guide**

**Interview guide (v2.0/27.11.2024)**

**Semi-structured interview guide**

Thank you for your time in taking part in this interview. We are interested in your perspective regarding the barriers and facilitators to referrals to specialist sleep services for suspected Obstructive Sleep Apnoea (OSA) and narcolepsy. There is no right or wrong answer, we are simply interested in your experience of treating patients with OSA and narcolepsy. We are also interested in learning about the factors that influence referrals to specialist sleep services.

**Questions**

1. How long have you been practising as a GP?

- *Are you working full-time or part-time?*
- *Tell me about your practice – how many GPs, how many sites, in which areas, etc.?*
- *How can a patient make an appointment? Any triage systems in place?*
- *Where do your patients live: urban rural areas?*

2. Was there a particular reason why you volunteered to participate in this study?

- *A particular interest in OSA or narcolepsy? Or a desire to expand your knowledge?*

3. Some of your patients may report feeling very sleepy during the day. Do you ask your patients how feeling very sleepy during the day *(*also called Excessive Daytime Sleepiness EDS*) has impacted on their quality of life? Do you have any concerns about it? (Automatic motivation)

- *Familiar with the Epworth Sleepiness Scale?* *Do you know where to access it?* (Psychological capability)
- *What other conditions might you consider when a patient reports EDS?* (Psychological capability)
- *When do you use and how do you interpret it?*
- *If a patient reports feeling very sleepy as the primary concern when booking an appointment, who would typically see the patient? A GP/ nurse?*
- *How do you differentiate between fatigue and excessive daytime sleepiness?*

**I am interested in learning about your experience of treating patients with OSA and narcolepsy. The first part of the interview will focus on OSA and the second on narcolepsy.*

**Obstructive sleep apnoea**

4. Could you describe your last patient with suspected OSA symptoms to me, what symptoms did he or she present with? (Psychological capability)
5. What happens when you encounter a patient presenting with the symptoms of OSA? Could you describe the consultation?

- *Any tools to help you diagnose? At what point would you use these tools? (Physical capability)*
- *Do you have enough time and resources to conduct a consultation? (Physical capability)*

6. Are there any preparatory or introductory steps at your practice to help you screen for OSA? *(Physical capability)*

- *Do you have a triage system in place? If yes, how does the triage system work?* (Automatic motivation)
- *Local or national guidelines? Regulations to encourage you to screen for OSA?*

7. What specialist sleep services are available in your area, if any? (Physical opportunity)

- *If not, how far is the nearest specialist service?*

8. Have you ever made a referral to specialist services for OSA? (Reflective motivation)

- *If yes, to what services? What was your experience of the referral process?* (+Physical capability)
- *Any barriers/facilitators?* (Physical opportunity)
- *If no, are you aware of the current referral process? Which services would you refer OSA patients to?* (Psychological capability)
- *Are the roles of sleep specialists & GPs clearly defined? Do you refer all OSA patients & at what point do you refer patients to a specialist?* (Psychological capability)

9. Would you know what are the waiting times for a patient with OSA to see a sleep specialist? (Physical opportunity)

- *Would some patients/carers consider seeking private healthcare due to lack of sleep services/long waiting times?* (Physical opportunity)
- *What is your view of private sleep laboratories? (Reflective motivation)*

10. I am interested in learning about the treatment for patients with OSA. Who would typically diagnose OSA? Who would typically provide treatment including prognosis to those patients? Would it be the specialist sleep service or other professional or you or do you share care for these patients? (Reflective motivation)

- *If treated by GPs: How well equipped do you feel to treat patients with OSA?*
- *Any care pathways/guidelines for OSA? If not, what information or resources would be useful?* (Reflective motivation)
- *Does continuity of care represent an issue?* (Reflective motivation)
- *What are your views of the available treatment for OSA? Do you perceive the treatment as beneficial to your patients?* (Reflective motivation)
- *What would you say are the barriers to OSA treatment?* (Reflective motivation)

11. Do you provide information to patients regarding OSA? Where is the information from? (Psychological capability)

**Narcolepsy**

12. Could you describe your last patient with suspected narcolepsy symptoms to me? (Psychological capability)

- **If cannot remember a patient -> go to next question*

13. What happens when you encounter a patient presenting with the symptoms of narcolepsy? Is it rare? Could you describe how the consultation might look like? Do you feel that you have enough time and resources to conduct a consultation? (Physical capability)

- *What symptoms would you consider to be representative of narcolepsy? Have you ever come across a patient diagnosed with narcolepsy?*
- *How severe would it be before you suggest intervention?*
- *What would do if a patient reported that they have narcolepsy?*

14. Have you ever made a referral to specialist services for narcolepsy? (Reflective motivation)

- *If yes, to what services did you refer? What was your experience of the referral process?* (Physical capability)
- *Any barriers/facilitators?* (Physical opportunity)
- *Do you refer all patients with a suspected narcolepsy? (Psychological capability)*
- *If no, are you aware of the current referral process to those services? Which services would you refer patients presenting with narcolepsy to? (Psychological capability)*

15. I am interested in learning about the treatment for patients with narcolepsy. Once patients are diagnosed with the condition, who would typically provide treatment to those patients? Would it be the specialist sleep service or you or do you share care for these patients? (Reflective motivation)

- *If treated by GPs: How well equipped do you feel to treat patients with narcolepsy?* (Social opportunity)
- *Any care pathways/guidelines for narcolepsy? If not, what information or resource would be useful?*
- *Are the roles of sleep specialists and GPs clearly defined?* How do you manage ongoing care of patients with narcolepsy? *(Psychological capability)*
- *Would the sleep service/specialist provide diagnosis & treatment?* (Physical opportunity)
- *Does continuity of care represent an issue?* (Reflective motivation)
- Could you tell me what treatments are available for narcolepsy? *What are your views of the available treatment for narcolepsy? Do you perceive them as beneficial to your patients?* (Reflective motivation)
- *What would you say are the barriers to treatment for patients with narcolepsy?* (Reflective motivation)

16. Would you know what are the waiting times for a patient with narcolepsy to see a specialist? (Physical opportunity)

- *Would you know whether any of your patients consider using private healthcare due to lack of sleep services/long waiting times/NHS initiative? What is your view of private sleep laboratories? (Reflective motivation)*

17. What are your views of sleep studies e.g. polysomnography? Do you think that your patients can benefit from having e.g. polysomnography? Do you find reports from sleep laboratories useful? (Reflective motivation)

- Can you refer patients for sleep studies without referral to a consultant? (Physical opportunity)

18. What training, if any, have you received on sleep disorders? (Reflective motivation)

- *What training do you think might be useful for GPs in this area (i.e. OSA/ narcolepsy)? Have you noticed areas where there might be a training deficit?*
- *Would you participate if it were offered? Would you have time to offer such training?*
- *Are you involved in any educational activities of future/junior GPs?*

19. If you lived in an ideal world, what resources would you need to facilitate referral to specialist sleep services for OSA or narcolepsy? (Physical opportunity + social opportunity)

Add participant information sheet and consent form?
